# Heart rate synchrony as an objective biomarker for therapy outcome in affective disorders

**DOI:** 10.1101/2024.08.29.24312787

**Authors:** Clara C. Gernert, Peter Falkai, Christine M. Falter-Wagner

**Author notes:** **Correspondence:** Clara C. Gernert, Christine M. Falter-Wagner.

## Abstract

The therapeutic alliance is widely recognized as a key predictor of psychotherapy outcome, though it is predominantly assessed through subjective self-reports. This proof-of-concept study investigated whether interpersonal movement and heart rate synchrony can serve as objective biomarkers for therapy effectiveness within naturalistic cognitive behavioral therapy (CBT) settings. Twelve patient-therapist dyads were analyzed in a short-term follow-up setting. Physiological signals were continuously recorded using wrist-worn wearable sensors, while head and body movements were captured via video recordings. After each session, both dyad members completed alliance questionnaires. Patients also provided standardized symptom ratings. Interpersonal synchrony was quantified using lagged-windowed cross-correlations across both modalities. Results confirmed the presence of synchrony in movement and heart rate during CBT sessions. Notably, in-phase heart rate synchrony measured in the initial session significantly predicted changes in both, patients’ self-rated global severity index and depressive symptom severity over time, independent of baseline symptom burden. Among subjective measures, only therapists’ ratings of the therapeutic alliance significantly predicted short-term therapy outcome. These findings highlight interpersonal physiological synchrony as a meaningful and unobtrusive objective marker of therapeutic engagement and effectiveness. Incorporating wearable biosensing into psychotherapy research might offer a scalable, objective approach to understanding and enhancing treatment effectiveness. Based on this proof-of-concept study, future studies with larger samples are warranted to refine synchrony-based predictive models and explore their potential for personalizing treatment choices for improving patient-therapist matching in evidence-based psychotherapy.

## 2 Introduction

In light of the high prevalence rates of mental health disorders (Kieling et al., 2024; Thom et al., 2024; GBD 2019 Mental Disorders Collaborators, 2022) - along with their substantial subjective burden, the risk of chronic progression and symptom aggravation when left untreated - there is a growing need to identify early, reliable predictors of psychotherapeutic outcomes. This need is further exacerbated by the limited availability of psychotherapy as a treatment resource. According to Grawe’s neuropsychological theory, mental disorders may be conceptualized as the result of persistent failures in the regulation of internal inconsistencies or motivational incongruence (Grawe, 2007, pp. 350-353). The primary therapeutic objective is thus not merely symptom relief, but the restoration of functional consistency, achieved through emotionally and cognitively corrective experiences embedded in the therapeutic process (Grawe, 2007, pp. 353-356). Psychotherapy fosters these experiential transformations to reduce inner conflict, thereby alleviating psychological distress and promoting well-being (Grawe, 2007, p. 353). Psychotherapy itself constitutes a complex bio-psycho-social system in which patterns of language, emotion, and cognition are continuously shaped and reshaped through reciprocal patient-therapist interactions (Gelo and Salvatore, 2016). The interaction itself is inherently dynamic, involving two interdependent systems, patient and therapist, which mutually influence and regulate each other in real-time, within the given asymmetric setting of psychotherapy (Strong, 1968). Within this relationship, the co-occurrence of interpersonal and emotional features plays a pivotal role in forming the therapeutic alliance, a construct widely acknowledged as critical for therapeutic effectiveness (e.g., Prusinski, 2024; Del Re et al., 2012, 2021; Flückiger et al., 2012, 2018, 2020; Wampold, 2015; Horvath et al., 2011; Martin et al., 2000; Horvath and Symonds, 1991). Originally conceptualized by Bordin, the TA is characterized by three interrelated components: agreement on goals, consensus on tasks, and the development of an emotional bond (Bordin, 1979). Importantly, Bordin emphasized that while therapeutic techniques and interventions are necessary tools, it is the TA that enables patients to engage with these methods in a meaningful way, fostering adherence and receptivity to change (Bordin, 1980, p.2). Alliance scores obtained during the early and terminal stages of therapy have been found to predict outcomes more accurately than those measured during intermediate phases (Adler, 1988; Horvath, 1981; Plotnicov, 1990). Furthermore, alliance ratings collected after the first session have been shown to predict premature termination (Kokotovic and Tracey, 1990; Plotnicov,1990), highlighting the prognostic value of early relational markers. However, concepts exist suggesting that therapeutic success is not only a result of working alliance quality per se but also depends on the individual characteristics of both therapist and patient (Saxler et al., 2024; Lindfors, 2021; Ackerman and Hilsenroth, 2003; Horvath and Luborsky, 1993; Mallinckrodt, 1991; Piper et al., 1991).

While the TA was traditionally framed as a product of cognitive-emotional engagement and verbal exchange, recent theoretical and empirical research has emphasized the embodied dimension of TA. Human interaction unfolds not only between minds but also between bodies. Interpersonal synchrony (IPS), defined as “the degree to which the behaviors in an interaction are non-random, patterned or synchronized in both timing [and] form.” (Bernieri and Rosenthal, 1991, p. 403) reflects one such embodied mechanism. IPS is not limited to non-verbal behavior, such as movement (e.g., Hoffmann et al., 2024; Zimmermann et al., 2021; Tsuchiya et al., 2020), but also manifests in speech (e.g., Amiriparian et al., 2019), physiology (Gernert et al., 2023; Behrens et al., 2020; Tschacher and Meier, 2019; Palumbo et al., 2017), and even brain activity (e.g., Dikker et al., 2014; Mu et al., 2016). Research has found IPS to be fundamental in fostering social relationships and cohesion by showing associations with rapport (e.g., Vacharkulksemsuk and Fredrickson, 2012; Miles et al., 2009), affiliation (e.g., Hove and Risen, 2009), successful cooperation (e.g., Behrens et al., 2020; Valdesolo and DeSteno, 2011), prosocial commitment (Mogan et al., 2017), and empathy (e.g., Marci et al., 2007). From a developmental perspective, biobehavioral synchrony has been proposed as a core mechanism for acquiring resilience (Feldman, 2020).

As an index of interpersonal connection, IPS might play an important role in psychotherapeutic treatment relationships (Wiltshire et al., 2020). While movement synchrony has been associated with therapy outcomes, physiological synchrony, such as heart rate (HR) or electrodermal activity, was mostly linked to the therapeutic alliance (Wiltshire et al., 2020) and empathy (Marci et al., 2007). Nevertheless, only a limited number of studies have examined physiological IPS in psychotherapy. Physiological synchrony variables like HR dynamics (Tschacher and Meier, 2019; Kodama, 2018), respiration (Tschacher and Meier, 2019), and skin conductance (Gernert et al., 2023; Marci et al., 2007) were assessed mostly in small samples and using conventional, lab-based monitoring systems. Although HR is a well-established biomarker for affective and cognitive states, and a key index of sympathetic and parasympathetic arousal (Coutinho et al., 2014), only a few studies have explored HR synchrony in therapeutic settings. Studies have primarily focused on its correlation with subjective ratings such as alliance (Tschacher and Meier, 2019) or empathy (Kodama et al., 2018), but less so on therapy outcome. While the TA gets often assessed by subjective reports, thereby introducing potential inconsistencies and biases, wearable-based biosensing technology offers a promising tool for capturing objective, continuous, and ecologically valid physiological data.

Building on the *In-Sync* model (Koole and Tschacher, 2016), which proposes that movement synchrony promotes interbrain coupling, affective co-regulation, and thereby therapeutic alliance, the current proof-of-concept study aimed to evaluate the presence and clinical relevance of HR synchrony in cognitive behavioral therapy (CBT). HR data from dyads of patients and respective therapists were continuously recorded using wearable biosensors during two naturalistic CBT sessions each. Concurrently, body and head movements were captured via a stationary video camera. Following each CBT session, patients and therapists completed standardized post-session reports on therapeutic alliance and perceived progress. Patients also reported their psychiatric symptoms and psychological distress using standardized questionnaires. HR and interpersonal movement synchrony were analyzed both as indices of TA and as potential predictors of therapy outcome, operationalized as a patient-reported change in symptom severity between the initial and follow-up CBT session. The study addressed four aims: (1) to confirm the emergence of IPS in heart rate during CBT sessions, (2) to investigate the association between IPS and symptom severity, as well as subjective alliance ratings, (3) to analyze whether IPS or subjective ratings predict therapy outcome, and (4) to assess whether patients’ initial depressive symptom severity might influence those associations.

## 3 Materials and methods

### 3.1 Participants

For the study, 25 patient-therapist dyads were recruited from the Clinic for Psychiatry and Psychotherapy (LMU University Hospital, Munich), encompassing both inpatient and outpatient departments. Participants with cardiac disease or implants, acute substance abuse, high-dose beta-blockers, acute neurological disorders, or severe skin defects were excluded from participation. All participants had either normal or corrected-to-normal vision. The therapists involved were not part of the research team and had no engagement in planning the study or analyzing the datasets. Out of those 25 dyads, 19 dyads participated in a short-term follow-up design, involving two CBT sessions per dyad, while six dyads completed only one session. From the 19 dyads in the follow-up group, four dyads were excluded due to technical failures during data acquisition in at least one of their two recorded sessions, resulting in incomplete datasets and subsequent removal from the analysis. Of the remaining 15 dyads, three dyads were excluded due to poor signal quality in the HR time series of at least one member of the dyad. After converting and preprocessing raw photoplethysmography (PPG) signals into HR time series, a quality check was performed (for details, see section 2.4.2.1), leading to the exclusion of these three dyads. As a result, 12 dyads with complete and analyzable data sets were retained for statistical analysis. At least two weeks passed between the initial CBT session and the follow-up CBT session for each of these 12 dyads.

The follow-up group comprised a mixed clinical patient sample (66.7% female, 33.3% male, M_age_ = 35.1 ± 13.8). Each patient had at least one (n = 8) or several (n = 4) diagnoses of a mental disorder according to ICD-10, Chapter V(F) (World Health Organization, 1993). The follow-up group included 8 different therapists (87.5% female, 12.5% male, M_age_ = 32.4 ± 8.6), with two therapists participating with two different patients and one therapist participating with three different patients. A full demographic summary is shown in Tables 1-3 (see the Supplementary Materials). All participants provided written informed consent, and the study received IRB approval from the LMU Medical Faculty Ethics Board (19-170).

### 3.2 Study design

The study was conducted in a designated therapy room at the Clinic for Psychiatry and Psychotherapy (LMU University Hospital, Munich). Each dyad member, patient and therapist, was equipped with a research-grade wrist-worn biosensor (*E4*, Empatica, Milan), worn on the non-dominant hand to minimize motion artifacts. Those wearable devices continuously recorded physiological parameters non-invasively throughout the CBT session. To capture head and body movements, a stationary video camera recorded the full course of each CBT session. Participants were seated in chairs positioned at approximately a 60-degree angle, facing each other. Verbal content and spoken language were neither recorded nor analyzed. At the beginning of each CBT session, therapists were instructed to set a timestamp on their wristband to facilitate the temporal alignment of the multimodal datasets. Subsequently, the dyad was left alone in the room for the following CBT session and received no specific instructions regarding spoken content. Session durations averaged M_duration_= 47.5 min (SD = 4.2 min), based on all recorded sessions (N_sessions_ = 24) across the follow-up sample (N_dyads_ = 12). Following each CBT session, both patients and therapists completed standardized post-session reports. Patients additionally rated their somatic and psychological symptom burden. A follow-up session was scheduled at least two weeks later, employing an identical study setup.

### 3.3 Materials

#### 3.3.1 Movement data

The CBT sessions were recorded using a standard video camera with a frame rate of 25 frames per second (fps). The entire session, excluding the initial two minutes, was used for assessing movement IPS, using the software *Motion Energy Analysis V3.10* (MEA) (Ramseyer, 2020). MEA is a frame-differencing method quantifying grayscale pixel changes in pre-defined regions of interest (ROI) (for more details see Ramseyer, 2020). In the current study, two distinct ROIs were defined, focusing on head and upper body movement.

#### 3.3.2 Heart rate

Empatica’s *E4* (Empatica, Milan) wristband sensor was used to continuously collect physiological data from both dyad members throughout the CBT sessions. During recording, data were streamed via Bluetooth to the *E4 Realtime* app and temporarily stored in the app’s cloud system. Following each CBT session, recorded data were transferred to the *E4 Connect* cloud platform, from which all raw datasets were subsequently downloaded and deleted afterward from the platform. Notably, the sensor collects only physiological signals without any personally identifiable information. No metadata was stored or transmitted, ensuring that E4 data cannot be traced back to individual participants. The wristband’s PPG sensor uses light signals at a sampling frequency of 64 Hz. Those light signals enable the extraction of cardiovascular dynamics, including HR, through pulse wave detection. For details on the preprocessing of PPG and HR signals, see section 2.4.2.1.

#### 3.3.3 Self-report questionnaires

After each CBT session, both patient and therapist completed the *Berner Post-Session Report 2000* (*BPSR-P 2000/BPSR-T 2000;* Flückiger et al., 2010). The BPSR-P comprises 22 items across eight subscales, while the BPSR-T consists of 27 items covering eleven dimensions. All items are rated on a seven-point bipolar Likert scale, ranging from −3 (*strongly disagree*) to +3 (*strongly agree*), with 0 indicating a neutral response. Following previous research (Ramseyer, 2019), we selected five patient-rated dimensions (i.e., *state alliance*, *trait alliance*, *progress*, *self-efficacy* (combining *mastery* and *clarification*), and *contentment of the bond*) and four therapist-rated dimensions (i.e., *state alliance*, *trait alliance*, *reactance*, and *progress*), to focus specifically on alliance- and process-related variables. Although Flückiger et al. (2010) originally conceptualized TA as a single dimension composed of three items in both questionnaires, we opted to differentiate this construct for analytical clarity. Specifically, we separated the three TA items into: a *state alliance* dimension (“Today I felt comfortable in the relationship with my therapist/client.” (in *BPSR-P*/*BPSR-T*)), and a *trait alliance* dimension (“The therapist/patient and I understand each other.” (in *BPSR-P/BPSR-T*), and “We are working toward shared goals.” (in *BPSR-T*); “I believe the therapist is genuinely interested in my well-being.” (in *BPSR-P*)). This decision was theoretically motivated by the distinct temporal focus of the items: while one targets the session-specific, momentary perception of the therapeutic bond (*state alliance*), the other two reflect a more generalized evaluation of the therapeutic relationship (*trait alliance*). For a detailed description of all selected dimensions with the belonging items, see Table 5 in the Supplementary Material.

Patients’ depressive symptomatology was assessed using the German version of the *Beck Depression Inventory – Second Edition* (BDI-II; Hautzinger et al., 2009). The BDI-II consists of 21 questions referring to the last 14 days, where each answer is scored with a value of 0 to 3. Higher total sum scores indicate a more severe depressive symptomatology. Patients’ overall symptom severity was evaluated using the German version of the *Brief Symptom Inventory* (BSI; Franke, 2000). The BSI measures psychological distress and clinically relevant somatic symptoms experienced over the past seven days on a four-point scale. The questionnaire consists of 53 items covering the following nine dimensions: somatization, obsession-compulsion, interpersonal sensitivity, depression, anxiety, hostility, phobic anxiety, paranoid ideation, and psychoticism. *Global Severity Index* (GSI) and *Positive Symptom Distress Index* (PSDI) are two key metrics from the BSI. While the GSI score is a strong indicator of the client’s overall psychological distress experience, the PSDI provides information about the intensity level of positively rated dimensions. In our follow-up sample, BDI, GSI, and PSDI scores were assessed after both sessions. For analyzing therapy outcome, we were particularly interested in the change of GSI (i.e., ΔGSI) and BDI (ΔBDI) over time. Positive values of both difference scores indicate a reduction of patients’ global symptom severity over time, while negative values indicate an increase. For descriptive statistics see Tables 6-11 in the Supplementary Material.

### 3.4 Data analysis

Data pre-processing and analysis of interpersonal physiological and movement synchrony were conducted using *R V4.4.0* (R Core Team, 2024) in *RStudio 2024.09.01.394* (Posit team, 2024) and *Python 3.13.3* (Python Software Foundation, 2024) in *Visual Studio Code*.

#### 3.4.1 Movement synchrony

##### 3.4.1.1 Preprocessing and synchrony calculation

To account for the non-stationarity of movement data and the potential temporal delays between dyadic responses, a lagged windowed cross-correlation analysis was employed for each dyad and both ROIs, as extracted from MEA. The analysis was conducted using the *rMEA* package (Kleinbub and Ramseyer, 2021). Raw MEA time series, sampled at 25 Hz, were initially imported. To improve signal quality, potential movement outliers were removed using the rMEA::MEAoutlier function. Outliers were defined as data points exceeding a dynamic threshold of ten times the standard deviation, applied in the positive direction. This way only unusually high movement values were considered as outliers and thus excluded from further analyses. Following outlier correction, all time series were standardized using rMEA::MEAscale to ensure comparability across dyads and sessions. Subsequently, lagged cross-correlations were computed using the rMEA::MEAccf function. The cross-correlation function (CCF) was calculated over rolling windows of 60 seconds with 30-second increments, using a symmetric lag range of ±5 seconds. This allowed the detection of delayed but temporally aligned patterns of movement between interactants within a specific time range. CCF values were calculated with non-absolute values, preserving the directionality of synchrony, and allowing us to differentiate in-phase synchrony from anti-phase synchrony. To enable statistical comparability, all CCF coefficients were Fisher z-transformed. To derive movement synchrony indices from the CCF matrices, a peak-picking algorithm was applied within each window. Two types of synchrony were computed per window: in-phase synchrony, based on all positive CCF values, and anti-phase synchrony, based on negative CCF values. For anti-phase synchrony, the strongest negative CCF value per window was identified by the maximum of the absolute negative CCF values. The negative sign was then reintroduced to preserve the directionality. For each dyad, peak values were aggregated across all analysis windows by calculating the mean for both synchrony types (i.e., in- and anti-phase), separately for both ROIs (i.e., head and body movement). This procedure yielded four movement based IPS metrics per dyad: in-phase head synchrony, anti-phase head synchrony, in-phase body synchrony, and anti-phase body synchrony.

##### 3.4.1.2 Control analysis

To ensure that the computed IPS indices capture genuine interpersonal movement coordination rather than statistical artifacts, a pseudosynchrony control analysis was conducted. Two surrogate data approaches were applied, described by Moulder et al. (2018): *data shuffling* and *section sliding.* These procedures were applied to the movement data from the initial session of the follow-up group. Following the analytical rationale of Plank et al. (2023), based on Moulder et al. (2018), we assessed (i) whether movement synchrony showed temporal structure across the session, and (ii) whether significant alignment could be observed within short time windows of the entire CBT session (i.e., 10-second segments). Pseudosynchrony metrics for both control approaches were computed using the same methodology and parameter settings as applied for real movement IPS calculation (i.e., lagged windowed cross-correlation and peak-picking). For each of the four IPS movement indices (i.e., in- and anti-phase IPS for head and body movements), Bayesian one-sample t-tests were performed to compare real synchrony scores against those pseudosynchrony scores, each based on 200 surrogate iterations per dyad. The results revealed strong to extreme evidence for genuine synchrony in all four real-dyad IPS movement indices. Paired t-tests further confirmed these effects (for all p < .001). Those results support both hypotheses (i) and (ii) (for details, see Table 13 in the Supplementary Material), validating the applied lagged cross-correlation approach as a robust and sensitive method for quantifying IPS based on MEA-derived motion energy in this sample.

#### 3.4.2 Heart rate synchrony

##### 3.4.2.1 Preprocessing IPS calculation

A common challenge in using wearable sensors are movement artifacts and typically lower sampling rates compared to standard laboratory-grade equipment. Motion artifacts can significantly compromise the quality of PPG signals, thereby impacting the reliability of stress detection at the individual level. It gets even more critical when analyzing interpersonal physiological synchrony.

To address this issue, the PPG time series from each participant were pre-processed using a protocol adapted from Campanella et al. (2024), who used physiological data from the *E4* (Empatica, Milan) sensor to evaluate stress reactions: First, raw PPG signals sampled at 64 Hz were segmented into 60-second windows with 15-second overlaps, corresponding to a final HR sampling frequency of 1/15 Hz. Each window was band-pass filtered using a Chebyshev Type II filter (0.5-5 Hz, order = 4, stopband attenuation = 20dB (Campanella et al., 2024; Liang et al., 2018)). Peaks in the filtered signal were identified with a minimum inter-peak distance of 0.4 seconds. Inter-beat intervals (IBI) were computed, with a constraint of IBIs to lie between 0.4 and 1.2 seconds. This constraint refers to the assumption of heart rate to be within the physiologically plausible range of 50 bpm to 150 bpm, especially when considering the setting of participants with no heart disease and a measurement while seated. IBI signals were then converted to a HR signal, measured as beats per minute (bpm). Subsequently, the resulting HR time series was cleaned using a relative deviation filter: if the percentage change between two consecutive HR values exceeded 25%, the latter value was marked as invalid. Such changes in HR within a brief time are considered unrealistic for HR data (Cheung, 1981). If fewer than 25% of values in the whole HR time series were missing, linear interpolation was applied. HR time series with more than 25% missing or corrected values were excluded from further analysis. To ensure the temporal plausibility of the HR signal, a second custom quality check was applied. For each HR time series, the relative change between each value and its counterpart two minutes prior was computed. If the deviation exceeded 25%, the value was flagged as physiologically implausible. Dyads in which either the patient’s or therapist’s pre-processed HR time series exhibited excessive deviations were excluded from the analysis. This resulted, as mentioned earlier, in the exclusion of three dyads (see section 2.1).

##### 3.4.2.2 Synchrony calculation

Similar to movement IPS calculation (see section 2.4.1.1), HR synchrony was calculated using lagged windowed cross-correlation. HR time series were first normalized across sessions and dyads by using the rMEA::MEAscale function. Cross-correlation matrices were computed using rMEA:MEAccf across 5-minute windows with 2-minute increments, and ±30-second lag range. Non-absolute CCF values were used for HR synchrony to distinguish the directionality of synchrony (i.e., in-phase versus anti-phase). All CCF coefficients were Fisher z-transformed for comparability. The peak-picking algorithm was applied to each CCF window. Afterward, all peaks per dyad were aggregated, leading to two HR synchrony indices per dyad: in-phase and anti-phase HR synchrony (for details about the peak-picking method, see section 2.4.1.1).

##### 3.4.2.3 Control analysis

To ensure that calculated HR synchrony reflected genuine interpersonal physiological coupling, a pseudosynchrony control analysis was performed using the same surrogate data procedures as described in section 2.4.1.2 (i.e., dyad-wise *data shuffling* and *section sliding*) (Moulder et al., 2018). Both approaches were applied to HR data from the initial session of the follow-up group. All surrogate datasets were processed using the same lagged windowed cross-correlation and peak-picking procedure as for real HR synchrony analysis (i.e., 5-min windows, 2-min increments, ±30-s lag). To evaluate (i) the temporal structure of HR synchrony, and (ii) synchrony within short-term segments (here defined as 1-minute intervals), Bayesian one-sample t-tests were performed comparing real dyad synchrony against pseudosynchrony distributions (n_permutation_= 200). The results revealed strong to extreme evidence for genuine synchrony in both real in-phase and anti-phase components across dyad shuffling (BF_10_ = 37.9 and 29.7, respectively) and segment shuffling (BF_10_ = 2143.9 and 2510.7). Paired t-tests further confirmed these effects (for all p < .002). Together, these results support the robustness of the applied calculation method for IPS in HR dynamics.

##### 3.4.2.4 Descriptives Statistics

A paired Wilcoxon signed-rank test revealed significant differences in individuals’ HR dynamics between patients and their respective therapists across both CBT sessions. In both sessions, patients exhibited significantly higher mean HR values than their therapists. In session 1 (S1), patients (P) had a mean HR_P_S1_ = 86.4 bpm (SD = 7.4) compared to therapists’ (T) mean HR_T_S1_ = 77.7 bpm (SD = 4.3), *W* = 75, *p* = .002. In session 2 (S2), the pattern remained consistent, with patients showing a mean HR_P_S2_ = 84.4 bpm (SD = 9.3) compared to therapists’ mean HR_T_S2_ = 75.8 bpm (SD = 4.7), *W* = 66, *p* = .034. A summary of the HR statistics can be found in Table 4 and Figure 1 (see both in the Supplementary Material).

**Figure 1.**
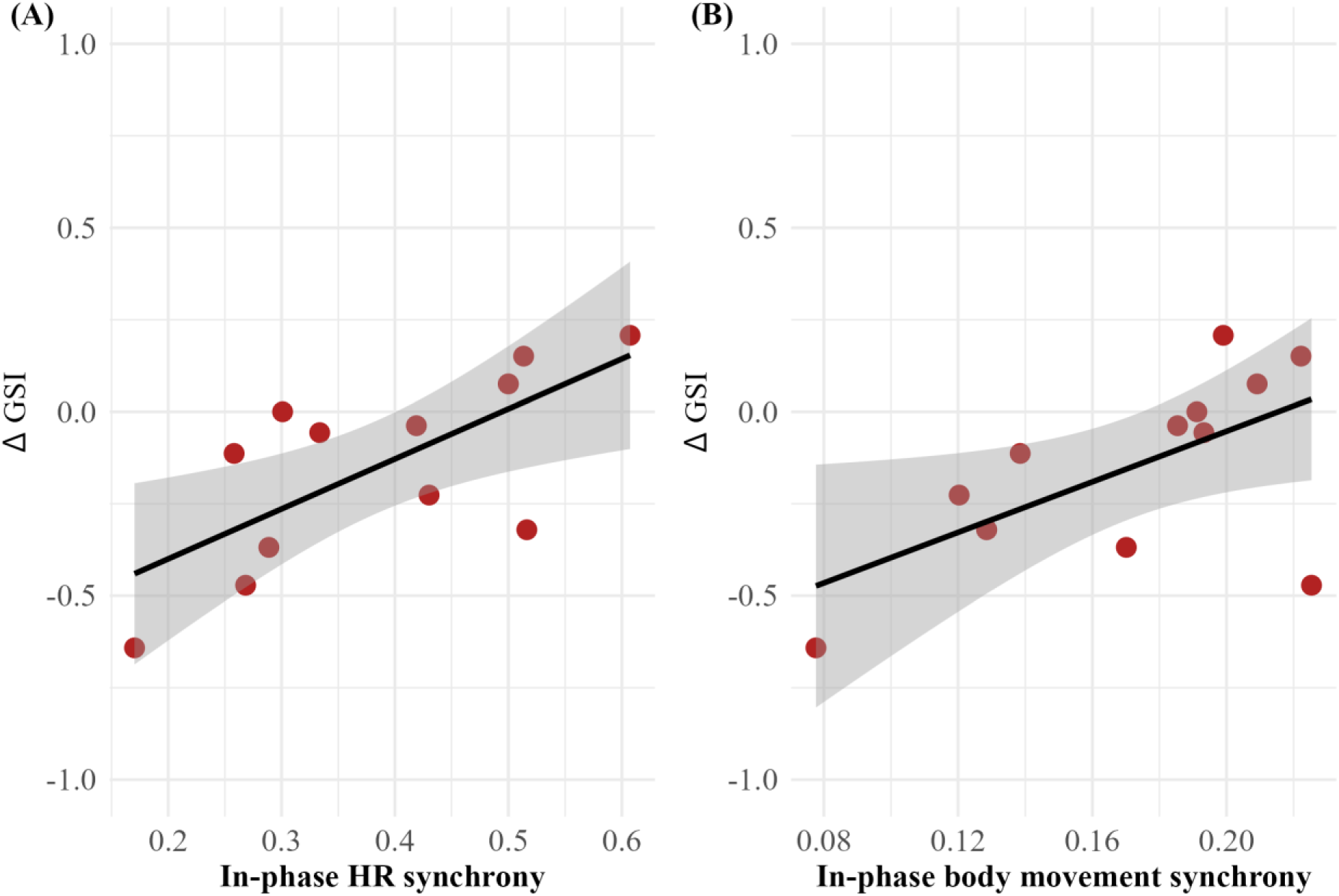
In-phase interpersonal synchrony as a predictor for symptom change. *Note.* Change in patients’ global severity index (ΔGSI) was predicted significantly by two in-phase synchrony indices. Positive ΔGSI values indicate a reduction in patients’ initial GSI over time, while negative ΔGSI values indicate an increase in GSI. A ΔGSI value of zero reflects no change in patients’ GSI ratings over time. A linear trendline and its 95% confidence interval (shaded area) illustrate the prediction of ΔGSI. Synchrony indices were calculated using the method of lagged windowed cross-correlation with peak picking (see section 2). Higher in-phase synchrony for both modalities during the initial CBT session was linked to a decrease in patients’ GSI over time, while lower in-phase synchrony indices were linked to an increase in symptomatology. (A) In-phase heart rate (HR) synchrony predicts ΔGSI, explaining 48% of the variance. (B) In-phase body synchrony also significantly predicted ΔGSI, explaining 37% of its variance.

#### 3.4.3 Statistical Analysis

Given that both HR and movement IPS scores significantly exceeded surrogate synchrony values, both modalities were considered in the following analyses, to (1) investigate the relationship between objective synchrony measures and subjective reports, including patients’ symptom severity and patients’ and therapists’ post-session reports, (2) examine whether IPS or subjective ratings are related to short-term therapy outcome, and (3) assess whether initial depressive symptom severity might influence those associations.

For the first part (1), correlation analyses were conducted using data from the initial session of the follow-up group: patients’ rated symptom severity scores (i.e., GSI and BDI), patients’ and therapists’ post-session reports, and HR and movement synchrony indices as objective variables.

For the second part (2), two outcome variables were included in the correlation analysis: ΔGSI, representing changes in patients’ general psychological distress experience, and ΔBDI, reflecting changes in depressive symptom severity over time. Variables that were significantly correlated with either one of the outcome variables were subsequently entered as predictors into a simple linear regression model. All linear regression models met the necessary statistical requirements, including the independence of residuals and homoscedasticity.

For the third analysis (3), patients were grouped into two subgroups based on their initial BDI-II scores, following the criteria of the BDI-II manual (Hautzinger et al., 2009): patients with scores within the range of moderate to severe depressive symptomatology (i.e., [20-63]) were classified as a *depressive* subgroup, while patients with scores in the range of minimal to mild depressive symptomatology (i.e., [0-19]) were classified as a *non-depressive* subgroup. Separate correlation analyses were conducted for both subgroups, based on the previously performed correlation matrices in (1) and (2).

## 4 Results

### 4.1 Interpersonal synchrony and post-session ratings

Patients’ ratings of *trait* and *state alliance* were positively associated with post-session ratings (*r* = .707, *p* = .010). Patients’ *trait alliance* ratings were further associated with patients’ perception of *progress* (*rho* = .815*, p* = .001) and *self-efficacy* (*rho* = .691, *p* = .013). Patients’ s*tate alliance* ratings also correlated with their perception of *progress* (*rho* = .652, *p* = .022), and *progress* ratings were strongly related to *self-efficacy* (*r* = .860, *p* < .001).

Therapists’ ratings of patient *reactance* were negatively associated with patients’ reports of *progress* (*r* = –.629, *p* = .028) and *self-efficacy* (*r* = –.609, *p* = .036). Therapists’ *reactance* scores also showed a significant negative association with their own ratings of *state alliance* (*rho* = –.641, *p* = .025).

No significant correlations were found between IPS indices and patients’ baseline symptom severity. However, patients’ *trait alliance* ratings were inversely associated with initial GSI (*rho* = –.729, *p* = .007). In-phase HR synchrony was positively associated with therapists’ *state alliance* ratings (*rho* = .651, *p* = .022). Notably, none of the patients’ post-session report dimensions showed significant association with any IPS variable. For a detailed summary of all correlations see Tables 14-18 in the Supplementary Material.

### 4.2 Prediction of symptom change

Simple linear regression models confirmed that three IPS variables significantly predicted ΔGSI: in-phase HR synchrony (*R*^2^ = .483, *F*(1,10) = 9.356, *p* = .012), anti-phase HR synchrony (*R*^2^ = .524, *F*(1,10) = 10.997, *p* = .008), and in-phase body synchrony (*R*^2^ = .367, *F*(1,10) = 5.801, *p* = .037). In addition, anti-phase HR synchrony significantly predicted ΔBDI (*R*^2^ = .355, *F*(1,10) = 5.493, *p* = .041). As shown in Figure 1, higher in-phase HR and body synchrony were associated with a greater reduction in psychological distress experience, while low synchrony scores were related to symptom increase. Conversely, higher anti-phase HR synchrony was associated with an increase in both GSI and BDI over time (see Figure 2).

**Figure 2.**
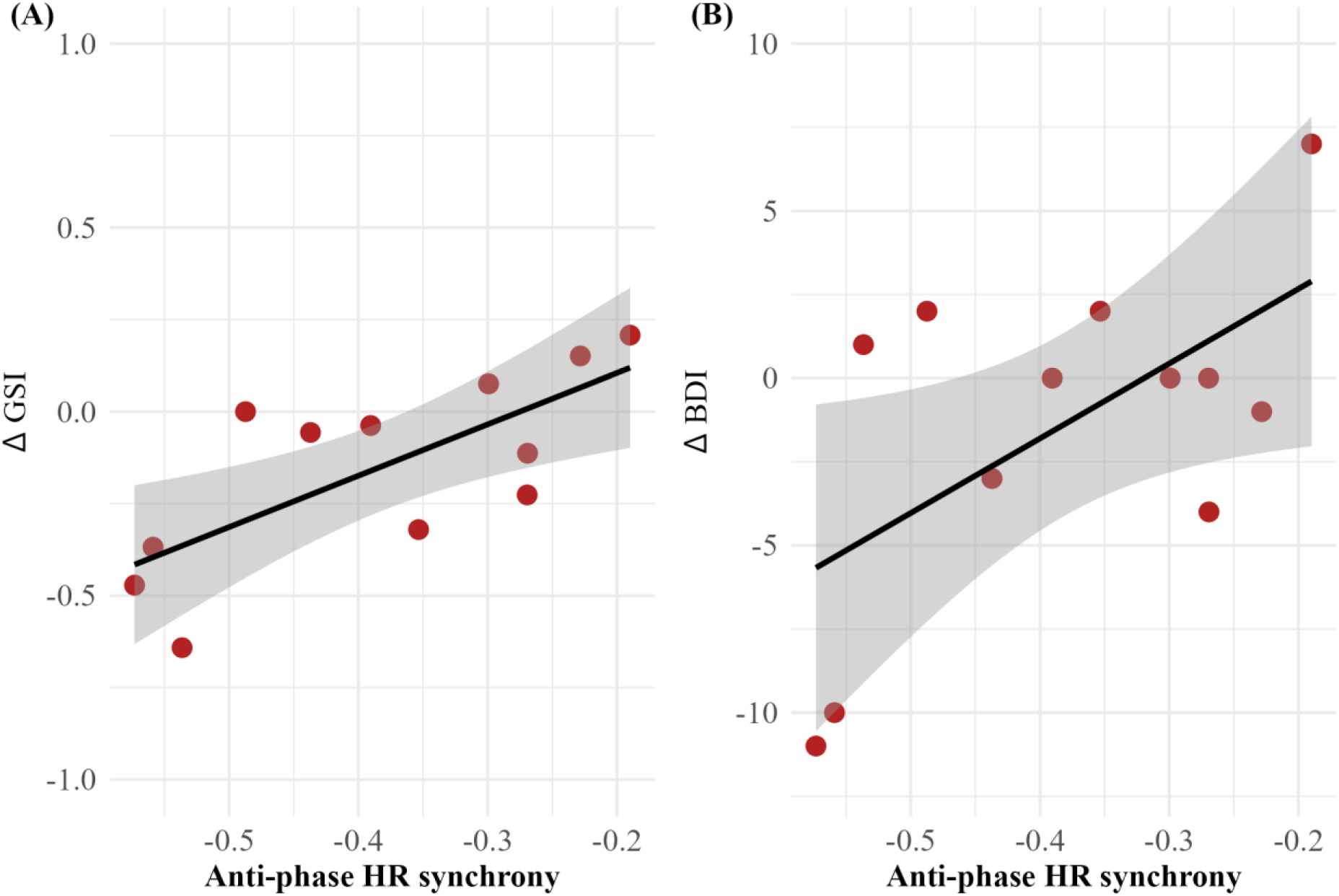
Anti-phase heart rate synchrony as a predictor for symptom change. *Note.* Anti-phase heart rate (HR) synchrony was a significant predictor for change in patients’ global severity index (ΔGSI) and for change in patients’ depressive symptom severity over time (ΔBDI). Positive ΔGSI and ΔBDI values indicate a reduction from patients’ initial GSI / BDI score over time, while negative ΔGSI and ΔBDI values indicate an increase in GSI / BDI. ΔGSI / ΔBDI values of zero reflect no change in patients’ GSI / BDI ratings over time. A linear trendline and its 95% confidence interval (shaded area) illustrate the prediction of both change variables. Greater anti-phase HR synchrony scores during the initial CBT session were linked to an increase in patients’ GSI and BDI scores over time, while lower anti-phase synchrony scores were linked to a decrease in symptomatology. (A) Anti-phase HR synchrony predicts ΔGSI, explaining 52% of the variance in ΔGSI. (B) Anti-phase HR synchrony was also a significant predictor for ΔBDI, explaining 36% of its variance.

Among the subjective post-session ratings, only therapists’ assessments of *state alliance* were significantly associated with both ΔGSI (*rho*= .609, *p* = .035) and ΔBDI (*rho* = .626, *p* = .029). However, when included in a linear regression model, therapists’ ratings significantly predicted only ΔBDI (*R*^2^ = .407, *F*(1,11) = 6.859, *p* = .026), not ΔGSI (p > .05).

None of the linear regression models revealed a significant interaction effect with patients’ initial symptom severity. An overview of the correlation matrices is provided in Tables 19-21, and a summary of all linear regression models is presented in Table 22 (see Supplementary Material).

### 4.3 Subgroup Analysis

To explore differences in IPS and outcome associations based on initial depressiveness, the follow-up sample (N = 12) was split into two subgroups: moderate to severely depressed (n = 6) and none or mildly depressed (n = 6), based on baseline BDI scores (see section 2.4.3).

#### 4.3.1 Moderate to severely depressed subgroup

In this subgroup, in-phase HR synchrony was very strongly associated with ΔBDI (r = .986, *p* < .001), and anti-phase HR synchrony also showed a significant association (r = .899, *p* = .008). These results suggest that higher in-phase and lower anti-phase HR synchrony were linked to greater reductions in depressive symptoms in depressive patients. Patients’ *state alliance* ratings were positively associated with ΔGSI (ρ = .878, *p* = .021), and *self-efficacy* ratings correlated significantly with ΔBDI (r = .872, *p* = .024). As within the entire follow-up sample, therapists’ *state alliance* ratings were significantly associated with ΔBDI (ρ = .939, *p* = .005).

#### 4.3.2 None to mildly depressed subgroup

Among patients with none or mild depressive symptoms, in-phase HR synchrony was significantly associated with ΔGSI (r = .832, *p* = .040). Patients’ ratings of *trait alliance* showed a significant positive association with ΔBDI (ρ = .912, *p* = .011), while *contentment with the bond* was significantly related to ΔGSI (r = .861, *p* = .028).

## 5 Discussion

Empathy is believed to foster the development of therapeutic alliance and is recognized as a robust predictor for therapeutic success (Horvath and Bedi, 2002). However, the conventional subjective assessment of therapeutic alliance overlooks the intricate intertwining of client and therapist through language, cognition, and emotions, their physiological states, and embodied communication. In the current study, we aimed to address this gap by incorporating physiological data collection alongside subjective reports from therapists and patients in a naturalistic CBT follow-up design. Wearable sensors continuously measured heart rate dynamics, while questionnaires were used to capture subjective ratings of therapeutic alliance and patients’ symptomatology.

An initial sanity check revealed significantly higher HR and movement IPS levels in real dyads compared to randomly generated surrogate dyads, highlighting that IPS existed as a non-random phenomenon when screening entire CBT sessions. This is in line with previous research reporting IPS in psychotherapy (Wiltshire et al., 2020). It also highlights that we were able to capture the construct of physiological IPS by wearable devices. In-phase HR synchrony correlated positively with therapists’ ratings of *state alliance*, aligning with prior findings that physiological synchrony relates to the concept of therapeutic alliance (Tschacher and Meier, 2019). In contrast, we did not find a significant correlation between patients’ and therapists’ post-session ratings of therapeutic alliance. This discrepancy underscores the occurrence of divergent subjective experiences among dyad members, despite their shared interaction. Such variability is common not only within the therapeutic context but also in daily interactions, reflecting the aspect of individuality in patterns of cognition, experiences, motivation, beliefs, and personality structure. In psychotherapy, where patients’ experiences take precedence over those of the therapist, reciprocity as well as mutual responsiveness regarding each other’s experiences and emotions are therefore limited. Patients’ mental health conditions could serve as another confounding variable. More than half of the patients included in our follow-up group were clinically diagnosed with an affective disorder, primarily depression. Depressive symptoms such as low mood, loss of interest, low energy, or lack of pleasure (see ICD-10, World Health Organization, 1993) can also contribute to difficulties in interaction (Kupferberg and Hasler, 2023). However, this insignificant finding underscores the limitations of the use of subjective ratings for predicting therapy success.

In-phase HR synchrony explained 48%, while anti-phase HR synchrony explained 52% of the variance in ΔGSI. Specifically, higher in-phase HR synchrony indices were associated with a greater reduction in symptom intensity, whereas high anti-phase and low in-phase HR synchrony indices were linked to an increase in symptom intensity levels over time. In contrast, among subjective measures, only therapists’ state alliance ratings significantly predicted symptom change, specifically the change in depressive symptom severity. We did not observe any significant interaction effects between patients’ initial symptom severity and the predictive value of IPS indices or subjective ratings in our linear regression models for short-term therapy outcome. Further research is necessary to analyze the causal relationship, including whether specific psychiatric symptoms are associated with decreased HR synchrony in interaction in general, whether this effect remains consistent across various interaction partners (i.e., therapists), and whether the level of HR synchrony increases in parallel with decreasing symptom burden over time.

While showing promising results concerning the use of objective biosensing of interpersonal dynamics, this proof-of-concept study is limited due to the relatively small sample size and exclusion of datasets with high artifact rates in physiological data. As we were interested in physiological IPS, low data quality from even one dyad member led to the exclusion of the whole dyad. Wearable sensors are usually more sensitive to motion artifacts, leading to lower data quality. While most physiological IPS research traditionally relied on classical devices like electrocardiograms for HR measurements, our study highlights the potential of wearable devices in capturing physiological dynamics during CBT. Those devices are less intrusive, which is important when aiming to capture naturalistic interaction, particularly in the setting of psychotherapy, where content is often quite intimate.

Additionally, the heterogeneity of our patient group might limit the generalizability of our findings. Nevertheless, among the participants in our study, over half presented with at least one diagnosis falling within the spectrum of affective disorders (ICD-10 F3 diagnosis; World Health Organization, 1993). This cohort is notably representative of patients commonly encountered in the setting of CBT. Given the frequent occurrence of comorbid diagnoses among psychiatric patients in clinical practice, our findings underscore the potential utility of physiological IPS as a transdiagnostic measure for therapy outcomes. This proof-of-concept study primarily aimed to explore the feasibility and potential of using wearable sensors to capture interpersonal physiological dynamics during CBT sessions in a real-world clinical setting. Based on the outcome of this proof-of-concept study and previous analyses (Riisager et al., 2025; Gernert et al., 2023; Schwartzmann et al., 2020) there is converging evidence of the potential of portable biosensors in psychotherapy research warranting future validation studies in larger sample sizes and preferably longitudinal designs. Although our follow-up assessment period was relatively short, spanning a minimum of two weeks, this timeframe was a pragmatic compromise, balancing the need for meaningful outcome assessment with the high patient turnover in a psychiatric clinic. Notably, prior work suggests that the predictive strength of the TA is not linearly related to time; rather, evidence suggests that alliance assessments taken early on or toward the end of treatment appear to offer more insight into treatment efficacy compared to ratings made during therapy (Horvath and Symonds, 1991). These findings support the idea that meaningful therapeutic change can indeed be captured within a relatively brief follow-up period. Nevertheless, despite the limited timeframe, the majority of our patients showed measurable changes in symptom severity. Due to the sample size, we could only run regression models with one predictor variable. Future studies with larger and diagnostically stratified samples, as well as extended follow-up periods, are needed to validate and extend these preliminary findings. By having a larger sample size, multivariate analysis should be calculated to control for confounders and interaction effects.

In general, a major challenge in the study of IPS lies in the heterogeneity of methodological and statistical approaches. This variability extends across multiple dimensions, including the type of signal analyzed (Gregorini et al., 2024; Ramseyer and Tschacher, 2014), as well as the specific segment of interaction selected for analysis. While some studies focus on the entire session (e.g., Zimmermann et al., 2021; Tschacher and Meier, 2019; Kodama et al., 2018), others limit their analysis to shorter specific time windows (e.g., Ramseyer and Tschacher, 2014). Further variation arises from the method of synchrony calculation and parameter settings (e.g., absolute vs. non-absolute values; global mean vs. mean of windows’ peaks; window size, lag, and increment). Moreover, approaches to generating surrogate data to assess pseudosynchrony vary, ranging from dyad shuffling (e.g., rMEA::shuffle function), to data shuffling and section sliding (e.g., Moulder et al., 2018). This heterogeneity not only hampers comparability across studies but can also lead to different results depending on the analytical strategy, as previously shown (e.g., Schoenherr et al. (2021); Luehof (2019); Altmann et al., 2022; Tschacher and Meier, 2019).

These challenges are also reflected in our work. In a previous study based on a different follow-up sample from the same overall dataset (Gernert et al., 2023), we analyzed EDA and movement synchrony during a fixed 10-minute segment at the beginning of the first session (excluding the first five minutes). Besides differences in the signal quality of EDA and HR, this segment-based approach allowed for the inclusion of dyads even in cases where technical issues occurred later in the session, whereas the present study required uninterrupted recordings for full-session analysis, resulting in different inclusion and exclusion criteria. Furthermore, the analytic frameworks differ for the calculation of movement IPS. In Gernert et al. (2023), we used absolute cross-correlation values and non-overlapping windows (following e.g., Tsuchiya et al., 2020; Ramseyer and Tschacher, 2014). The present study builds upon previous HR synchrony studies (e.g., Tschacher and Meier, 2019, Kodama et al., 2018) where the entire session was assessed for IPS calculation. Thus, movement IPS was also considered for the entire CBT session. We used non-absolute cross-correlations to distinguish between in-phase and anti-phase synchrony and applied overlapping windows. The previous study relied on dyad-shuffling (rMEA::shuffle function) to create surrogate data (Gernert et al., 2023), while the current analysis applied data shuffling and section sliding as recommended for small sample sizes (Moulder et al., 2018). While both approaches are justified within their respective analytic frameworks, the differences in parameter settings and inclusion criteria likely contributed to heterogeneous results for movement IPS. Notably, when analyzing a 10-minute segment of a CBT session, significant IPS above pseudosynchrony was found solely for head movement (Gernert et al., 2023). IPS calculations using the entire CBT session, as applied in the present analysis, showed significance above pseudosynchrony scores for both head and upper body movement.

This underscores how methodological heterogeneity can impact results and highlights the need for greater methodological transparency and cross-validation of analytic approaches to ensure the robustness and comparability of IPS findings across studies.

In conclusion, our study demonstrates the value of utilizing wearable sensors in CBT settings. It emphasizes the predictive strength of HR synchrony as an objective biomarker for therapy outcomes. Moreover, integrating the concept of embodiment into further research on evidence-based psychotherapy might have the potential to tailor therapy approaches and enhance patient-therapist matching.

## Data Availability

The participants of this study did not give written consent for their data to be shared publicly, so due to the sensitive nature of the research supporting data is not available.

## 8 Author Contributions

All authors conceptualized the study. C.F.W. secured research funding. C.C.G. recruited participants and collected data. C.C.G. and C.F.W. developed the analysis plan. C.C.G. pre-processed and analyzed the data. C.C.G and C.F.W. wrote the first draft of the manuscript. P.F. and C.F.W. supervised the study. All authors approved the manuscript.

## 10 Acknowledgements

We are thankful for patients’ and therapists’ commitment and contribution to our study. C.C.G. gratefully acknowledges E. P. for her support and encouragement on a personal level.

